# The use of generative artificial intelligence applications by undergraduate dental students

**DOI:** 10.64898/2026.05.25.26353910

**Authors:** Mario Brondani, Jonathan Rafael Garbim, Shimae Soheilipour, Vincent Lee

## Abstract

**Background:** Higher education has been transformed by the rapid integration of generative artificial intelligence (GenAI) tools into academia. The objective of the present study was to examine how and for what purposes senior undergraduate dental students use GenAI tools in academic assignments.

**Methods:** This cross-sectional study uses data from three written assignments submitted by two consecutive cohorts of graduating fourth-year dental students at the Faculty of Dentistry at the University of British Columbia, for a total of 120 students. The assignments focused on different subjects where students had to offer their views, including community water fluoridation. When using GenAI, students were asked to disclose whether and how such tools were used, and for what purpose. Descriptive statistics (e.g., means, frequencies, and proportions) were conducted via IBM SPSS Statistics (Version 27.0).

**Results:** From the two cohort of students, 102 (85%) disclosed the use of GenAI tools in at least one assignment; of these, 69 (67.6%) reported using these tools in all three assignments. ChatGPT was by far the most frequently used GenAI tool, reported by 89 students (87.2%). Nine students (8.8%) did not specify which tool they had used. The majority of the students (91.2%, n = 93) reported using GenAI for proofreading or grammatical editing. About 9.8% of the students (n = 10) reported more substantive uses, such as relying on GenAI to generate in part or in full the assignment, and/or assessing the credibility of references.

**Conclusions:** In our study, the use of GenAI tools was highly prevalent among senior undergraduate dental students for editorial purposes. A smaller but notable proportion of students engaged in more substantive uses that may carry academic and ethical risks. There is a need for structured AI literacy training and clear, dentistry-specific guidelines to promote responsible and transparent use while safeguarding critical thinking, academic integrity, and professional judgment in dental education.

## Introduction

The adoption of artificial intelligence (AI) applications in higher education^1,2,3^ and research^2,4,5^ has seen unprecedented growth. Generative AI (GenAI) in particular, used for creating content such as text and images, is being rapidly adopted. GenAI uses large language models (LLMs) and training data to produce novel, human-like outputs. LLMs such as Chat Generative Pre-training Transformer (ChatGPT, OpenAI Incorporated®), Google’s Gemini (Google® DeepMind), Perplexity (Perplexity AI, Inc.), Copilot (Microsoft 2025) and Claude (Anthropic, AI Chatbot Desktop) have gained substantial momentum and continue to expand rapidly. Among these tools, ChatGPT, released free-of-charge on November 30, 2022, has experienced widespread adoption. ChatGPT has reportedly reached more than 815 million active users, defined as individuals accessing the platform at least once a week, and more than 5.8 billion monthly visits as of February 2026.^6^

ChatGPT enables users to ask questions on virtually any topic, request both general and specialized information, perform qualitative and quantitative data analyses, generate written content, and engage in dialogue. ChatGPT works through a conversational prompting interface. The material generated by GenAI can span a wide range of domains,^7,8,9^ including for academic purposes for almost 20% of its users.^6^ Consequently, a growing body of literature has examined the potential applications of ChatGPT and similar LLMs in education, research, and clinical practice.^10,11,12,13,14^

In terms of research, ChatGPT has been reported to assist with tasks such as improving scientific writing, supporting literature reviews, and facilitating data analyses.^10,12^ These benefits are accompanied, however, by notable limitations, including the risk of generating inaccurate information, fictitious references, and content that may lack rigor or verifiability as presented by Ashraf and Ashfaq (2024)^15^ and others.^16^

In terms of education, GenAI tools have used to support personalized learning and student engagement. At the same time, their utilization poses concerns related to ethics, transparency, bias, plagiarism, lack of originality, and the validity and reliability of AI-generated content.^12,15^ Some scholars have cautioned that excessive reliance on LLMs may have unintended consequences for students’ learning and well-being, including reductions in critical thinking skills,^17^ increased digital fatigue, technostress, diminished face-to-face interactions, and weakened interpersonal skills and emotional intelligence.^18^ Additional concerns including social isolation, loneliness, and anxiety have also been raised^19^ when GenAI tools are used to replace human interaction and independent cognitive effort.

Much of the empirical research on the use of LLMs in higher education to date has focused on medical education.^3,8,20,21^ On the other hand, the extent to which these technologies are being used in dental education is still emerging.^10,22,23^ And among the studies examining GenAI use by university students, it is possible that the prevalence and depth of LLM usage are underestimated.^24^

In light of these gaps, this studied examined how senior undergraduate dental students integrate GenAI tools into written academic work. Particular attention was placed on the purposes and depth of use disclosed by the students. It is important to understanding these patterns of use to get a better insight into how LLMs are shaping academic work in dental education within the broader context of the rapid expansion of GenAI technologies.

## Methods

This descriptive cross-sectional study examined undergraduate dental students’ self-disclosed use of GenAI tools in written academic assignments, and received approval from the University of British Columbia’s Behavioural Research Ethics Board (H25-01322). The self-disclosure was part of three written assignments submitted by two consecutive cohorts of graduating fourth-year dental students at the Faculty of Dentistry at the University of British Columbia (FoD-UBC): 57 students in 2024–2025, and 63 in 2025–2026, for a total of 120 participants. These assignments were part of a mandatory curricular course^25,26,27^ and were evaluated using a standardized grading rubric to ensure consistency across cohorts.

All assignments were submitted electronically through the university’s Canvas® platform. The assignments required students to critically reflect and articulate their perspectives on the following issues: (1) universal oral health care, with particular attention to issues of equality and equity in access to care among equity-seeking communities; (2) the health implications of sugary beverage vending machines in an inner-city school while considering the potential economic benefits generated to support the schoolchildren; and (3) the implementation of community water fluoridation while weighing in the benefits of fluoride against arguments raised by the anti-fluoridation movement.

The position chosen by the student on any of the issues presented could be any, although emphasis was placed on the quality of reasoning, critical analysis, and the appropriate use of evidence to support or challenge their arguments. Students were instructed to cite up to five references and to limit each assignment to a maximum of 1,000 words each, excluding the title page and references.

The course director informed the students that they were permitted to use any GenAI tools in the preparation and development of their assignments. They were required to disclose whether and how such tools were used (e.g., editing, idea generation, content development, reference-related assistance, and so on). Assurance was given to the students that the use or non-use of GenAI tools would not influence the grading of their work, but no specific guidance was provided regarding best practices for the use of GenAI in the academic context described herein.

Descriptive statistics (e.g., means, frequencies, and proportions) were used while inferential statistics (e.g., odds ratios) did not lead to any significant patterns of LLM use across assignments and cohorts, and are not shown. Data analyses were conducted using IBM SPSS Statistics (Version 27.0).

## Results

From all the 120 FoD-UBC students enrolled during the two-year study period, 102 students (85%) disclosed the use of LLMs/GenAI tools in at least one assignment. From these, 48 students did so in 2024–2025 and 54 students in 2025–2026. A total of 69 (67.5%) reported using LLMs/GenAI tools in all three assignments (32 students in 2024–2025 and 45 students in 2025–2026).

In terms of GenAI used, ChatGPT was by far the first option for 89 students (87.3%), followed by Perplexity, which was used by three students (2.9%), and Microsoft Copilot, which was used by one student (0.9%). Nine students (8.8%) did not specify which LLM/GenAI tool they had used.

The vast majority (91.2%, n = 93) of the 102 students who disclosed using LLMs/GenAI tools at least once used these tools primarily for proofreading or grammatical editing. A smaller proportion of students (9.8%, n = 10) reported more substantive uses, such as asking the tools to generate content related to the specific issue, summarize the evidence behind a chosen idea, explore the advantages and disadvantages of specific options, and identify and access the credibility of such references.

Some of the disclaimers are compiled below (Table 1), and were reproduced verbatim without edits. Based on the reported purpose of use, disclosures were grouped into three broad categories: editorial support (e.g., proofreading and grammatical editing), idea generation and content development (e.g., partial or full summarization, interpretation, or drafting assistance), and reference-related support (e.g., identification and/or appraisal of references). The list provided is not exhaustive as many students used AI for the same purpose, and the disclaimers they wrote were repetitive..

**Table 1:** Examples of GenAI use as disclosed by senior undergraduate dental students at the FoD-UBC.

|  | **Universal oral health care assignment** | **School vending machines Assignment** | **Community water fluoridation assignment** |
| --- | --- | --- | --- |
| **Use for editorial purposes** | "I used ChatGPT to re-word select sentences so that the language would flow better and sound more professional" (2025-26). | "Chat GPT was used to check for grammar and make sentences more concise" (2024-25). | "I used Copilot to help refine the wording and sentence structure of my own content" (2025-26). |
| **Use for generating ideas or content** | "I used ChatGPT to draft the content of this essay and to make sure the assumptions made were credible. I then added my own views on the topic." (2024-25) | "Perplexity was used for source finding and summarizing relevant key points. Rough letter outline was also provided by Perplexity" (2025-26) | "AI (ChatGPT) was used to seek clarification on what the role we can play in advocating, and to outline the evidence on my report" (2024-25) |
| **Use for referencing** | "AI was used to create the reference list to support the arguments made" (2025-26)) | "I used open AI to find the references, and organize them in citation" (2024-25 | "I acknowledge that AI (ChatGPT) was used to gather resources for this assignment" (2025-26) |

## Discussion

In this study, the purposes for which senior undergraduate dental students used GenAI tools to develop written academic assignments was assessed. The use of GenAI by at least 85% of the participants was higher than previous studies.^**Error! Bookmark not defined**.,28^

Such higher use attests for the increasing integration and normalization of LLM/GenAI tools within higher education.^10^ In response to this normalization, researchers and academics have called for the formal integration of AI literacy and training into academic curricula to maximize the educational benefits of these tools in dental education,^28,29^ and to position the profession at the forefront of technological innovation.^30,31^ But additional research is needed to inform evidence-based implementation and academic and curricular strategies around the use of these tools.^10,17,32,33^ The high prevalence of use observed in the present study suggests that GenAI tools are becoming embedded in students’ academic workflow.

As found elseswhere,^23^ ChatGPT (OpenAI Incorporated®) was the most frequently used LLM. Other tools such as Perplexity (Perplexity AI, Inc.) and Copilot (Microsoft 2025) were also used. The accuracy and reliability of LLMs in health-related academic contexts remain variable and task-dependent, with conflicting results.^34^ For example, a study examining embryology-related questions by Bolgova et al (2025) found stronger performance by ChatGPT and Claude,^35^ while research on histological questions reported no statistically significant differences among Google Gemini, Claude, Microsoft Copilot, ChatGPT, and DeepSeek.^36^ Although comparable evaluations were not performed in this study, and are only beginning to emerge within dental education,^37^ researchers consistently emphasize the need for critical verification of AI-generated information.^38^ It is important to verify the information generated to identify biases, quantify inaccuracies, and ensure that LLMs are used as supplementary educational tools rather than as replacement for instructor expertise or student critical judgment.^37,39^

In alignment with prior research, the majority of students reported using these tools primarily for proofreading and grammatical editing, as also observed by Ganjavi and colleagues (2024)^24^ and others.^40,41^ However, the finding that nearly one in ten students (9.8%) reported more substantive uses, such as asking the AI tool to develop the assignment, explore the advantages and disadvantages of specific options or assess the credibility of references, raises important concerns. The distinction between editorial assistance and cognitive delegation may become increasingly blurred when students rely on GenAI tools. Specifically, it is unclear whether these students fully appreciate the potential risks associated with LLM use, including exposure to inaccurate information, fabricated references, or content lacking rigor and verifiability.^11,16,15^ Furthermore, the 67.6% of students who reported using LLMs consistently across all three assignments may be at risk of overreliance, with unintended consequences for learning, critical thinking, and student well-being.^17,18,19^ Such patterns may reflect not only technological adoption, but also increasing reliance on AI-mediated cognitive support during academic reasoning processes. There is an urgent need for the development of dentistry-specific guidelines that promote best practices for academic AI use while explicitly fostering critical thinking and reflective judgment.^42^

Moreover, the boundaries between acceptable assistance and inappropriate delegation of academic work may become increasingly difficult to define when AI tools contribute to argument development, evidence appraisal, or reflective positioning. This distinction may be especially important in reflective and opinion-based assignments, where the educational objective extends beyond the production of text and includes the development of independent reasoning, professional judgment, and critical reflection.

Despite the findings presented herein, this study has limitations, including the fact that it relied on students’ self-disclosed use of GenAI tools and did not independently verify the extent or accuracy of the reported use. Even though students were made aware that the use or non-use of GenAI tools would not affect grading, some might have underreported the frequency or extent of their use, or chose not to disclose such use. Moreover, the data included a relatively robust number of assignments from two cohorts of students, but participants were from a single dental school, which limits generalization. Also, the assignments analyzed were primarily reflective or opinion-based and did not focus on clinical diagnosis, treatment planning, or prognostic decision-making. As such, it remains unknown whether these students use LLMs/GenAI tools in other academic contexts, for clinical decision support, or for exam preparation. More research is needed to explore how teaching strategies and assessment methods can evolve to preserve reflective learning, critical thinking, and professional judgment.

## Conclusions

The use of GenAI tolls is highly prevalent among senior undergraduate dental students. It seems to be increasingly embedded within academic assignments involving reflective and opinion-based writing. While most students are using these tools for editorial support, a notable proportion reported more substantive uses involving idea generation and evidence appraisal that may carry academic and ethical risks. Dental education programs need to move beyond restrictive or purely technical approaches to AI use and instead foster AI literacy, critical thinking, reflective judgment, and transparent academic practices. Such approach is important to preserve independent reasoning and professional development. These findings highlight the need for structured AI literacy training to foster responsible and transparency.

## Data Availability

All data produced in the present work are contained in the manuscript

## Acknowledgments

The authors declare no conflict of interest. Dr Brondani contributed to the conception and design of this manuscript, acquisition of the data, as well as drafting and critically revising the manuscript. Drs Garbim and Lee contributed to drafting, critically revising the manuscript and helped with the interpretation of the textual data as well as with the literature. Dr Soheilipour critically revised the manuscript and updated the references. All authors contributed to the final revision of the manuscript and formatting. The authors disclose the use of ChatGPT (https://chat.openai.com/) exclusively for language editing, including grammar, formatting, and clarity. No artificial intelligence–generated content (AIGC) tools or other large language models (LLMs) were used in the development of the manuscript’s content. This study did not receive direct project funding. However, Garbim received scholarship support from Fundação de Amparo à Pesquisa do Estado de São Paulo (FAPESP) (Grant No. 2022/04054-7). The funding agency had no role in the study design, data interpretation, or decision to submit the manuscript for publication.

